# The challenge of medication adherence to reduce cardiovascular risk in primary care: a mixed design multi-center study in underserved populations

**DOI:** 10.1101/2023.03.30.23287988

**Authors:** K Puschel, K Gonzalez, J Varas, J Sateler, H Aravena, D Greig, G Escalona, M Palominos, A Rioseco, A Cea, B Thompson

## Abstract

**Background:** Cardiovascular disease is a leading cause of death in Latin America. Internationally, low medication adherence is associated with 15% to 40% of excess cardiovascular deaths. In Latin America, the magnitude of low medication adherence and the factors associated with it, are not well known, especially among socially vulnerable populations. The aim of this study is to estimate the magnitude and associated factors of low medication adherence in a socially vulnerable population with high cardiovascular risk in Chile.

**Methods:** The study is based on a mixed-methods design. It included a multicenter cross-sectional design of a randomly selected clinical population of 900 participants, and a qualitative design based on the analytical framework model, that included patients and health team members, from three primary care clinics in Chile.

**Results:** Only 24.6% from the 886 (out of 900) patients who completed the study had “high” medication adherence, 24.9% had “regular,” and 50.4% had “low” adherence. Depression was the main factor associated with regular and low adherence combined (OR: 2.12; 95%CI:1.55-2.89). Confusion and tiredness were identified as barriers for adherence. Main facilitators reported by patients included better understanding of the medications, and availability of reminders. Clearer information and family support were identified by team members as initiators for improving adherence.

**Conclusion:** Low medication adherence is highly prevalent among patients with high cardiovascular risk in a low-income population in Chile. Quantitatively, depression was a significant risk factor for regular and low adherence; qualitatively, confusion and tiredness were identified as barriers. Clearer information and family support are identified as potential facilitators.

**What is new?:**

▪ Cardiovascular disease is a growing epidemic in Latin America
▪ Low medication adherence is a significant risk factor for cardiovascular disease

**What are de clinical implications?:**

▪ Low medication adherence is highly prevalent in socially vulnerable populations of high cardiovascular risk in Chile.
▪ Depression increased the risk of low medication adherence by more than two times in high-risk populations and is associated with the perception of confusion and tiredness in this group of patients.
▪ Primary care teams recognize the relevance of providing clearer information, reminder systems and developing family support strategies for improving medication adherence.

## Introduction

Cardiovascular disease (CVD) persists as the leading cause of death in the Americas^1^. The slow progress in reducing CVD over time, and the social inequality of disease consequences across populations is concerning^2,3^. In Latin America, there is a growing epidemic of cardiovascular disease representing 33.7% of total mortality in the region^4^. In Chile, cardiovascular disease is the leading cause of death(4) and 51% of the Chilean population is at moderate or high risk of having a cardiovascular event during the next ten years(6). Chilean populations of lower socioeconomic status have between 1.5 to 2 times higher risk of dying from cardiovascular disease compared to the population of higher socioeconomic status^7^.

Medication therapy can significantly reduce the morbidity and mortality from cardiovascular disease; however, its effectiveness is reduced if adherence to medication is low. Multiple studies, primarily from high income countries (e.g., United States, Canada, Japan, Australia, United Kingdom) have found that only about 50% of the population with cardiovascular risk has good medication adherence^8,9^. Low medication adherence is associated with 19% to 40% increased risk of experiencing a fatal or a non-fatal cardiovascular event in patients with moderate (i.e.10%-19% risk of developing CVD in the next 10 years) or high CVD risk (i.e. 20% or higher risk of developing CVD in the next 10 years) according to the traditional Framingham risk score ^8-11^.

Information concerning medication adherence in Latin America is scarce. In a meta-analysis of medication adherence recently conducted by Liu et al (2021)^9^, the authors included 46 articles but none of them were from Latin America. Primary care has been identified as a key setting for improving cardiovascular disease management in Latin America^12^. Primary care is the first point of contact for the majority of the population in Latin America^13^. It is where preventive strategies are developed (e.g., smoking cessation counseling, screening for high blood pressure, treatment for diabetes) and where prescriptions for chronic diseases are delivered. It is also the setting where most patients get tertiary prevention after returning from the hospital for cardiovascular events such as a myocardial infarction or stroke. Therefore, even small changes in primary care practices can have a big impact in cardiovascular health at a population level. In Chile, about 75% of the population seeks care in the public health care network. This primary care network manages the national cardiovascular program and offers free medication and clinical follow-up. In spite of this, 51.5% of the adult Chilean population is at moderate or high cardiovascular risk based on the Framingham risk score, according to the Chilean National Health Survey^6^.

Low medication adherence has been suggested as one factor to explain the limited success in cardiovascular risk management in primary care in Chile^12^. A number of reasons have been associated with medication adherence. They include patient-related factors, socioeconomic factors, health care system factors, therapy, and condition-related factors^14^. A better understanding of the magnitude of low medication adherence and the factors that influence it in primary care settings is essential to design effective interventions and improve cardiovascular care. The objective of this study is to examine primary care settings that include patients with high cardiovascular disease risk and to estimate the magnitude of non-adherence in such patients. Further, we will identify barriers and facilitators to improve medication adherence in such high-risk patients.

## Methods

### Study design and setting

This study is based on a mixed-methods approach that includes a multicenter cross-sectional quantitative study, and a qualitative approach based on a theoretical framework conducted in three primary care clinics in Chile. Primary care serves about 70% of the population in Chile and provides free care to the population including health promotion initiatives (e.g., physical activity programs), preventive care (e.g., smoking cessation counseling or diabetes screening) and therapeutic services (e.g. hypertension, coronary heart disease therapy). Entry into the national health program requires individuals to register at a Primary Health Care Clinic (PHC) where a multidisciplinary team of physicians, nurses, psychologists, and social workers among others, provides a number of health-related services organized into different programs. The cardiovascular program includes patients with hypertension, diabetes, coronary heart disease and cerebrovascular disease. National guidelines regulate the activities and assure resources for the cardiovascular program at the PHCs. As an example, individuals with risk factors for or with clinical CVD receive nutritional and nurse advice, psychosocial assessment, medical check-ups, lab testing (e.g., blood tests, EKG) and referral to specialty care if there is a need for specific tests (e.g., echocardiography testing). In addition, patients receive free medications as needed, such as angiotensin II receptor blockers, statins, metformin, or insulin.

This study was conducted in three PHCs in three different regions of the country. The clinics are located in La Pintana, Santiago (LP), San Clemente, Talca (SC) and Chiguayante, Concepción (Ch). The Clinic in La Pintana serves an urban population of 22,000 individuals of very low socioeconomic status. San Clemente and Chiguayante are semi-rural communities of low socioeconomic status located about 250 km and 500 km south of Santiago and serve a population of about 24,000 each.

### Participants

A random sample of 900 people (300 per PHC) was selected from a total population of 3000 patients (1000 per PHC) between 35 to 65 years old with moderate to high CVD risk registered at the three participating clinics. The selective criteria included participants with a moderate to high cardiovascular risk according to the Framingham score adapted to the Chilean population^15^, and that had been checked at the clinic during the last 6 months.

### Qualitative design

The qualitative design was based on the framework method^16^, which is based on the analysis of data gathered from a predefined set of themes and categories This method has been increasingly used in clinical and educational research^17,18^. The framework method approach was used to explore the experiences, barriers, and facilitators of a diverse group of participants between 35 to 65 years with cardiovascular risk; a group of health care team members also participated in focus groups. A snowball technique was used to select a total of 48 participants: 24 patients with high cardiovascular risk, and 24 primary care team members. The latter group included 5 physicians, 7, nurses, 3 nurse assistants, 2 pharmacists, 2 nutritionists, 2 psychologists, 2 physical therapists, and 1 social worker.

Focus group meetings were iterative, and subgroups of participants were invited to participate in three sequential rounds to obtain rich data of the specific dimensions of concern as described above.

### Instruments and procedures

For the quantitative data, personal interviews by phone were conducted with participants using a structured questionnaire of 35 questions that contained five attributes: demographics, medication adherence, psychosocial conditions (e.g., education level, income level, depression, anxiety disorders), risk factors and chronic health conditions (e.g., smoking, alcohol abuse, hypertension, diabetes) and clinical disease (e.g., coronary heart disease, stroke, kidney failure, cancer). Medication adherence was assessed using the Adherence to Refill and Medication Scale (ARMS). The ARMS questionnaire has been validated for populations with multi-morbidity and low-literacy levels. It is especially useful in primary care settings as it has high internal consistency (Cronbach’s alpha = 0.828)^19^. The scale has been validated in Spanish^20^ and showed high association with clinical variables such as blood pressure control and hospitalization for acute cardiac event^19,21^. High adherence is necessary for cardiovascular control, and different cut off points have been used in the ARMS instrument to define it^20^. We followed the cut-off point used by Lomper K. et al. (2018)^22^ and defined high adherence for a score of 12 in the ARMS instrument. However, we also analyzed a regular category for scores between 13 and 14 and a low adherence category for scores of 15 or higher. For the final analysis we combined regular and low scores to reflect fewer ideal scenarios. A written consent was sent from the authors of the ARMS instrument to the principal researcher of this study (personal communication with S. Kripilani, Dec 3^rd^, 2019). Psychosocial factors included depression and disorders related to alcohol abuse. Depression was assessed using the Whooley screening test that has been extensively used in primary care for the early detection of depressive disorders; it has high accuracy and low variability between countries^23^. Alcohol abuse disorders were identified using the Audit-C^24^ questionnaire that is included in the Chilean national guideline for detecting drinking problems in primary care populations. Multimorbidity was defined as the co-occurrence of two or more diseases^25^ and for polypharmacy we used the criteria most commonly reported in the literature that is, the use of five or more medications daily^26^.

Electronic chart records at the clinics were also systematically assessed to obtain additional information on participants risk factors (i.e., smoking, hypertension, diabetes, dyslipidemia) clinical conditions (i.e., blood pressure control, HA1c levels, coronary heart disease, stroke, kidney failure, cardiac heart failure, cancer, depressive disorders) and pharmacological therapy (type and number of medications).

For the qualitative aspect of the study, a semi-structured questionnaire was designed to explore experiences, barriers and facilitators of patients and health care team members concerning medication adherence. The questionnaire contained five key open-ended questions. All focus groups were conducted by two researchers (KG, HA or JS) in virtual mode given the COVID-19 pandemic restrictions during the study. They were audio-video recorded and fully transcribed for subsequent analysis.

### Analysis

The data were collected and registered in the Research Electronic Data Capture (RedCap) platform. Descriptive and analytic statistics were conducted using IBM-SPSS-28 software, professional version. (IBM-SPSS statistics)

Qualitative content analysis was conducted using Atlas.ti 9. Emerging codes and categories were identified and grouped into several themes in an iterative process conducted by two researchers (KG, KP). The researchers separately coded each focus group, met to compare codes, and discussed discrepancies until consensus was reached.

### Ethics

The project was reviewed and approved by the Institutional Review Boards at the Faculty of Medicine Pontificia Universidad Católica de Chile as well as Servicio de Salud Concepción and Servicio de Salud del Maule, Chile (CI200117003). This project is funded by the Chilean National Agency for Research and Development (ANID) (SA20I0001)

## Results

### Quantitative Analysis

The response rate of participants was 98.4% (886/900). The participating population consisted of Mestizos (38.9%), Whites (31.8%), Amerindians (14.8%) and a few Blacks (0.8%). Some 13.7% of participants did not report their race/ethnicity. The participants had low educational level with less than half having completed a secondary education, and very low or low-income levels. (Table 1). About 72% of participants had high cardiovascular risk and over half of them had hypertension and/or Type 2 diabetes. About 25% of the participants had clinical cardiovascular disease such as myocardial infarction (14.7%), stroke (14.0%) cardiac heart failure (15.6%) or kidney failure (11.5%). In addition, 12% of this population, with predominantly high cardiovascular risk, also had cancer at some time. Depression also was highly prevalent in this population (55.7%).

**Table 1.**
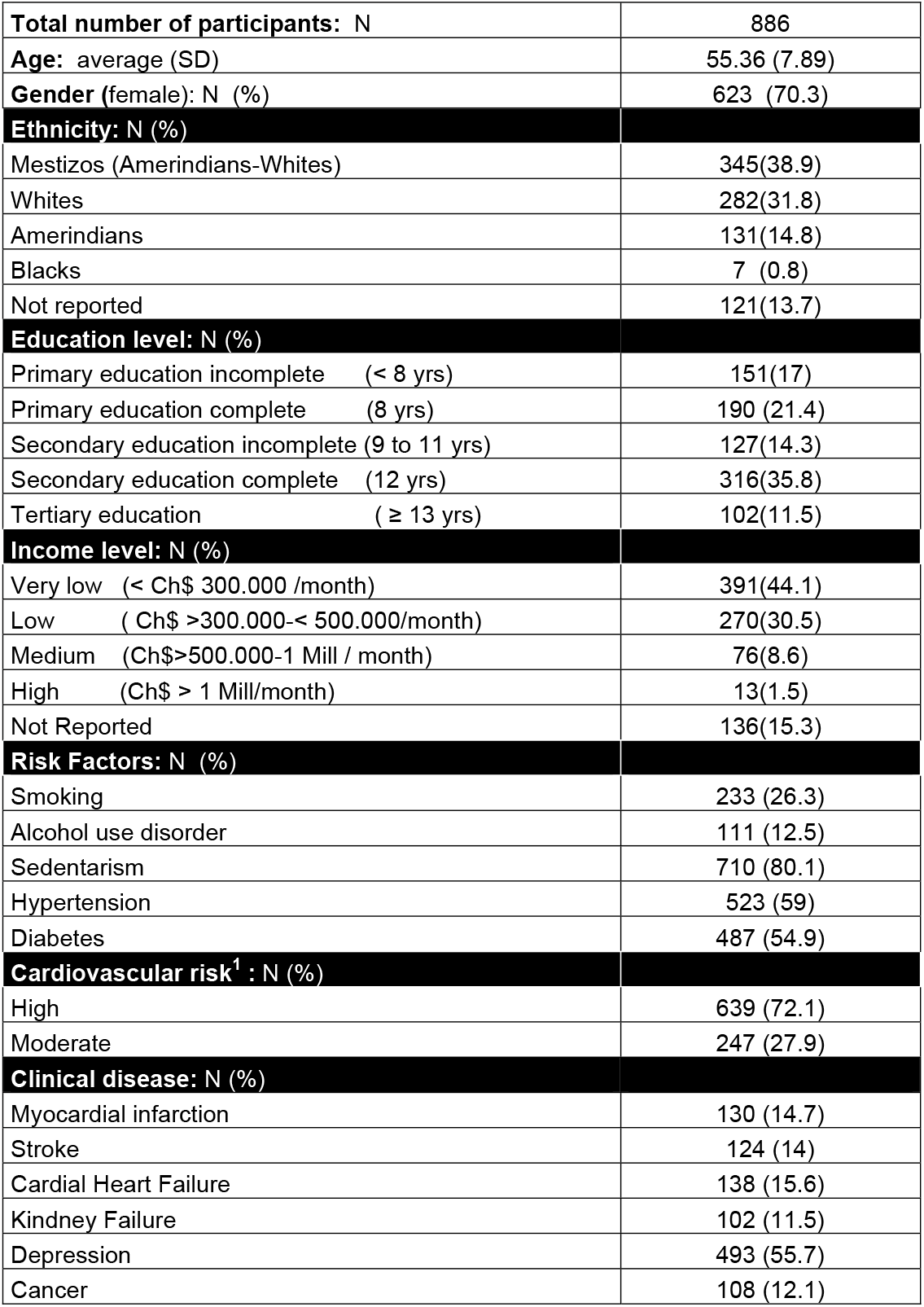
Demographic and clinical characteristics of participants.

Table 2 shows participants’ medication adherence as well as associated risk factors. Only 24.6% of this high-risk population achieved high medication adherence (ARMS 12 or less). The factors with the highest association for lower medication adherence (either regular according to ARMS (13 or 14) or low according to ARMS (15 or higher)), were very low-income level (OR = 1.46 95% CI: 1.05-2.04) and depression (OR = 2.12 95%CI: 1.55-2.89). Other factors such as ethnicity, multi-morbidity and polypharmacy were not significantly associated with lower medication adherence. The best multivariate model to explain low medication adherence included low socioeconomic status and depression (OR=2.09 95%CI: 1.21-3.58) (data not shown).

**Table 2.**
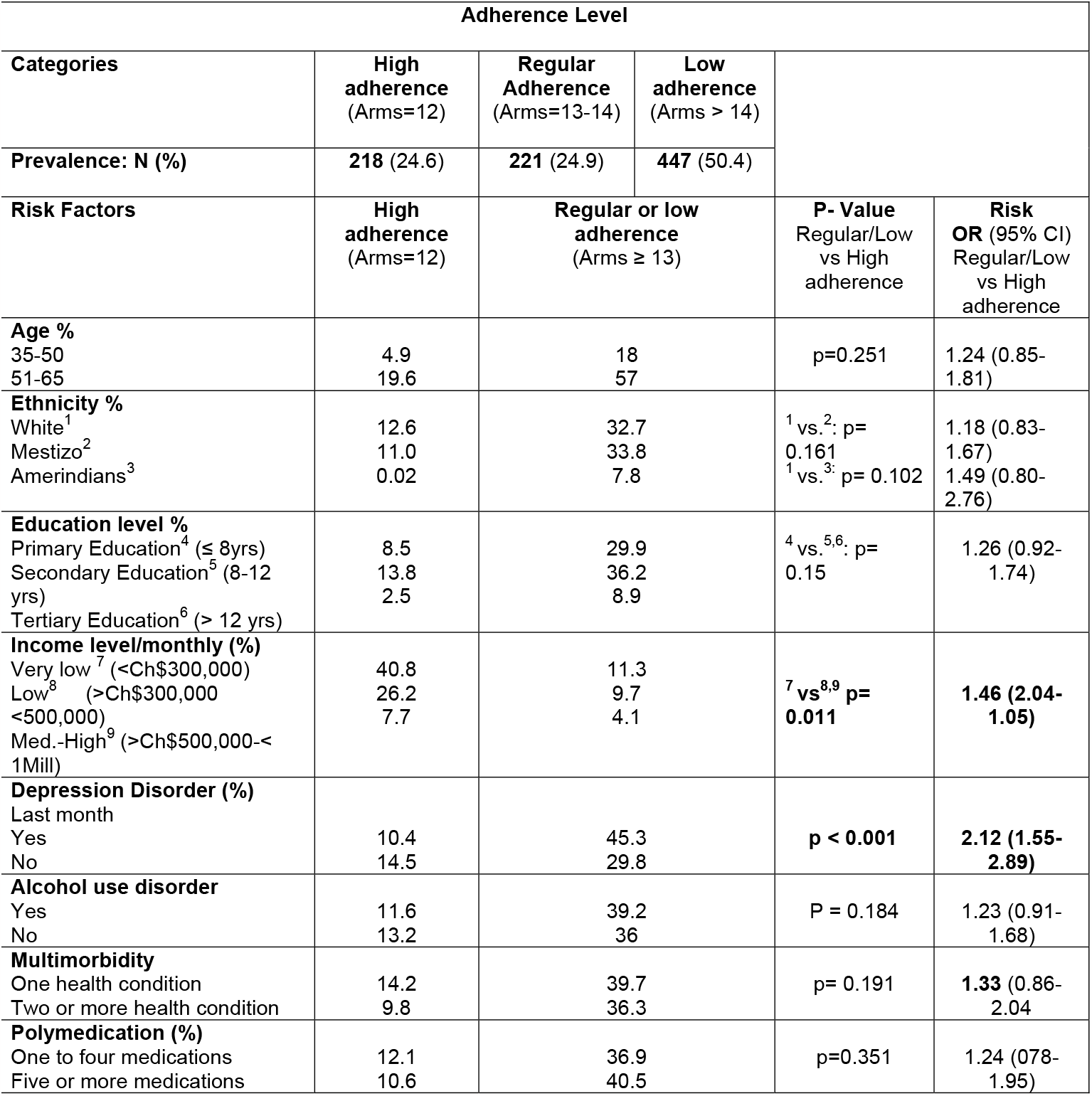
Medication adherence prevalence and related factors.

### Qualitative Analysis

Thirteen focus groups delivered in three rounds were conducted: nine with patients and four with team members. The total number of participants was 48 (24 patients and 24 team members) and the average number of participants per focus group was 5 (3-7). The average time per focus group was 64 minutes (58-72)

Figure 1 summarizes the main barriers and facilitators for medication adherence that emerged from the focus groups. There were differences in both the barriers and facilitators identified by patients compared to health care team members. *Confusion* was the main barrier identified by patients followed by *tiredness* and *misinformation*. In contrast, the main facilitator mentioned by this group was *understanding; that is, it was important* to comprehend the uses and actions of medications. R*eminders* and *nearness* were also mentioned, meaning it was important to have medication available and in sight.

**Figure 1.**
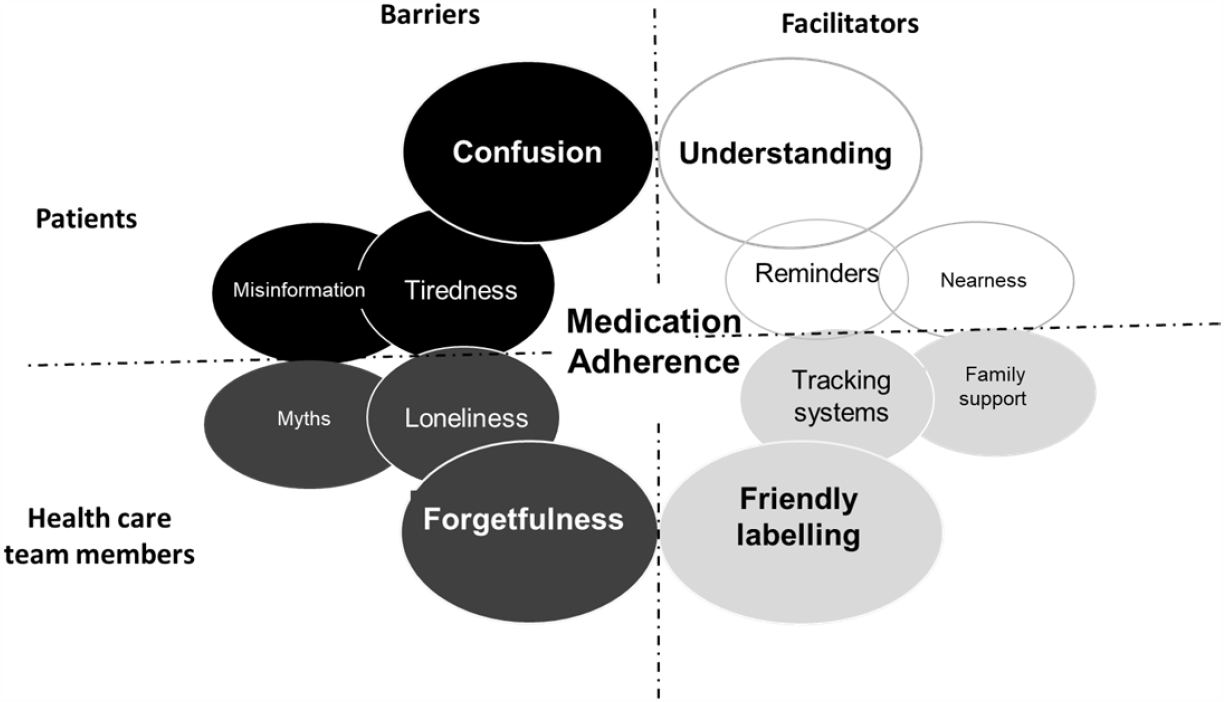
Factors associated with medication non-adherence by dimensions

Health team members mentioned *forgetfulness* as the main barrier for low medication adherence followed by *isolation* and *myths*, which were erroneous beliefs originating from lack of accessible or accurate information. *Friendly labelling, tracking systems* and *family support* were identified as main facilitators for improving medication adherence by the health care team members. Table 3 presents the themes that emerged during the focus group discussion and provides descriptive examples for each of them.

**Table 3.**
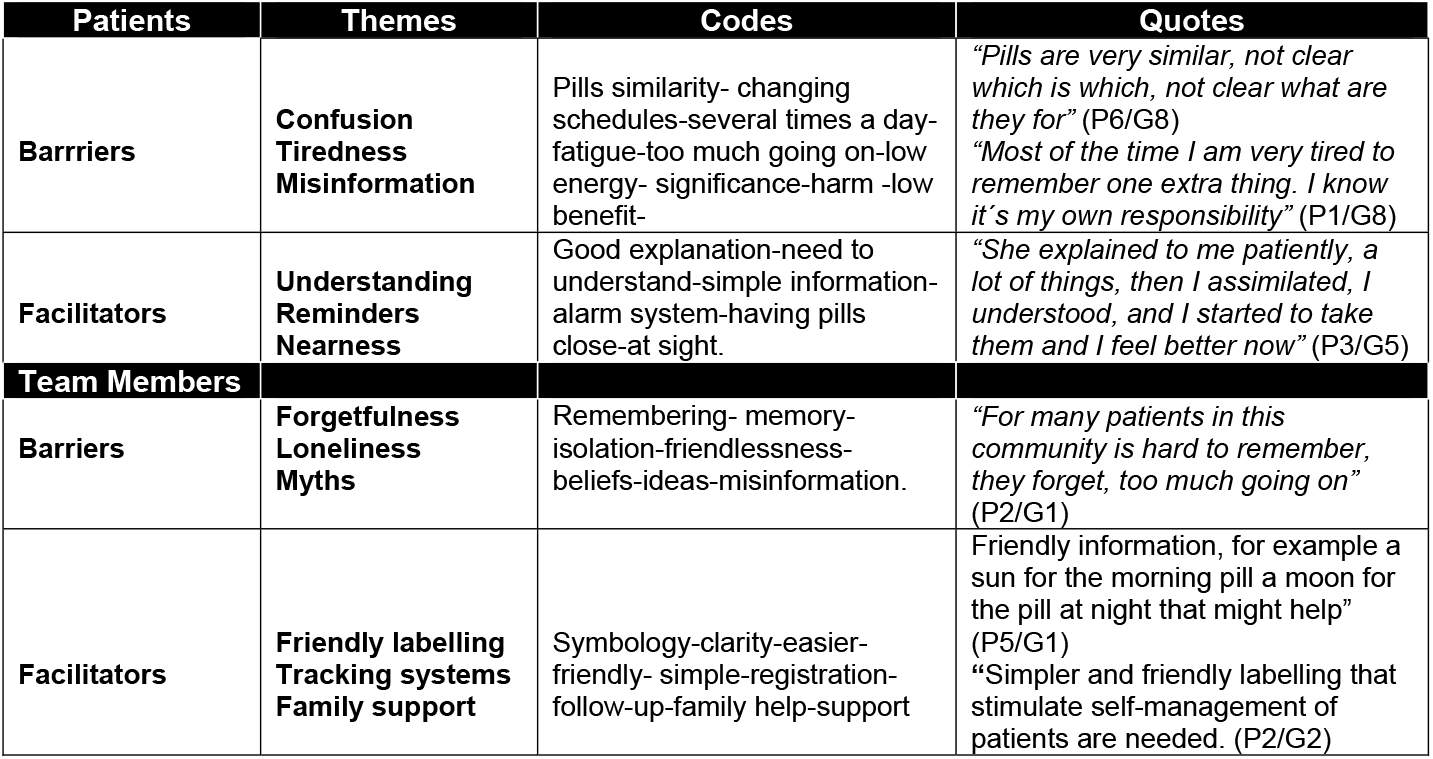
Barriers and facilitators associated with medication adherence.

## Discussion

This study indicates that low medication adherence is highly prevalent in a high-risk cardiovascular population in a Latin American population of low socioeconomic status. The low medication adherence prevalence of 75.4% reported in this study is higher than the average of 54% observed in the meta-analysis conducted by Liu et al (2021)^9^ that included North American, Western European, and Asian populations at cardiovascular risk. In the meta-analysis different instruments and cut-off points were included to estimate medication adherence. Our study falls in the high range of low medication adherence reported.

Low socioeconomic status was significantly associated with low medication adherence in our study. This association has also been described in many studies and systematic reviews^27,28^. However, the magnitude of the effect reported is highly variable. A number of mediator variables including psychosocial factors have been suggested to explain the variability observed in these studies^29,30^. Depressive disorders, for example, have been associated with low medication adherence in patients at cardiovascular risk in other studies^31^. Our results are consistent with that evidence and demonstrate that patients with depressive disorder have more than a two times higher risk of low medication adherence than patients with no depressive disorder. The magnitude of this effect is very similar to that published by Rasmussen et al (2007)^32^. Depression disorders were present in about half of our study population and were similar to the level found in the meta-analysis published by Khatib R, et al (2014)^33^. The high prevalence of depression found in our population and the significant association observed between depression and medication adherence might explain, in part, the high prevalence of low medication adherence of our population.

The synergistic effect of low socioeconomic status and cardiovascular disease has been described by Lee et al.^28^ in a 10-year cohort study that included more than 1.6 million hypertensive patients; they showed that this group had 2.46 times higher risk of cardiovascular deaths compared to the high income-good medication adherence group. On the other hand, there is consistent evidence showing that depressive patients have about 30% excess risk of cardiovascular death compared to non-depressed patients^34^. In the multivariate analysis of our study, we observed that the population of low socioeconomic status as well as depression had more than twice the higher risk for low medication adherence. This group of patients of low socioeconomic status and depression clearly represent a population that needs a unique approach for cardiovascular management at the primary care level. Organizational changes such as extra time for visits, additional psychosocial support interventions or different medication labelling could be pertinent strategies to implement in this group to achieve better outcomes and improve clinical equity^35^.

The qualitative approach integrated in our study, allowed us to explore in depth the association between patient and organizationally related medication adherence factors. Confusion and understanding were mentioned by patients as important factors that affected medication adherence. Both factors denoted the lack of clarity with which this population receives the recommendations provided by health team members, and also the difficulties to comprehend the significance of a process that needs to be performed every day and sometimes more than once a day. Health team members emphasized the relevance of delivering simpler, friendlier, and more homogenous messages to patients with chronic diseases at the primary care level.

Consistent with the high prevalence of depressive disorders found in this population study, focus groups participants identified tiredness as an important barrier for medication adherence. Tiredness has been identified as an expression of fatigue and is more prevalent among patients with chronic diseases living in socially deprived areas and with higher levels of isolation^36,37^. This profile represents well the characteristics of our primary care population. Loneliness was identified by health team members as a barrier for medication adherence and, in contrast, family support was mentioned as a facilitator for improving medication adherence. There is some evidence that shows that multidisciplinary family-based interventions are appropriate and might be effective for reducing cardiovascular risk in limited resources countries^38,39^.

This study has some limitations important to address. Participants were randomly selected from a clinical population of patients registered at their primary care clinic who have attended the clinic for evaluation during the past six months. Therefore, they represent an active population that might not be typical of the general population of patients with cardiovascular disease. However, the sampling was consistent with the main goal of the study that was to explore medication adherence in a real primary care population of patients with high cardiovascular risk and find potential factors that health care teams might use to improve their adherence. The cross-sectional design of the study limits the possibility of finding predictive factors associated with low adherence; however, the quantitative data obtained was integrated with qualitative information provided by patients with high cardiovascular risk and health team members. This allowed us to explore more comprehensive factors associated with low medication adherence.

## Conclusion

This is one of few studies conducted on cardiovascular disease and medication adherence in Latin America. This study shows that low medication adherence is highly prevalent in primary care patients with high cardiovascular risk in Chile. Socioeconomic variables as well as affective factors such as depression were associated with a higher risk of low medication adherence. Confusion and tiredness were main barriers mentioned by patients as barriers for improving medication adherence. Understanding the significance of their treatment and having a reminder system available were identified by patients as facilitators. Health team members highlighted the relevance of providing clearer information, and to identify a family member to support medication adherence among socially vulnerable patients.

## Data Availability

All data referred to in the manuscript is availability.

https://drive.google.com/drive/folders/1WC46ZQ5OP2QdRZ26lRGsTn4ojmInv7Pw

## Notes

### Competing Interest Statement

The authors have declared no competing interest.

### Clinical Trial

Cllinicaltrials.gov Prococol Registration NCT05395806

### Author Declarations

The project was reviewed and approved by the Institutional Review Boards at the Faculty of Medicine Pontificia Universidad Católica de Chile as well as Servicio de Salud Concepción and Servicio de Salud del Maule, Chile (CI200117003).

## References

(1) Lloyd-Sherlock P, Ebrahim S, Martinez R, et al. Reducing the cardiovascular disease burden for people of all ages in the Americas region: analysis of mortality data, 2000-15. Lancet Glob Health. 2019; 7(5): 604–612. doi: 10.1016/S2214-109X(19)30069-5

(2) Serón P, Lanas F. The Americas: a region that ages with disparity. Lancet Glob Health. 2019; 7(5): 540–541. doi: 10.1016/S2214-109X(20)30520-9

(3) World Health Organization. (2018). Time to deliver: report of the WHO Independent high-level commission on noncommunicable diseases. World Health Organization. Available at https://apps.who.int/iris/handle/10665/272710.[Accessed 25 November 2022]

(4) Lanas F, Seron P, Lanas A. Cardiovascular disease in Latin America: the growing epidemic. Prog Cardiovasc Dis. 2014; 57(3):262–7. doi: 10.1016/j.pcad.2014.07.007

(5) Bächler, R, Icaza G, Soto A, et al. Epidemiología de las muertes prematuras en Chile en la década 2001-2010. Rev. Med. Chil. 2017; 145, 319–326. doi: 10.4067/S0034-98872017000300005

(6) Ministerio de Salud. Informe Encuesta Nacional de Salud 2016-2017: Santiago de Chile; 2018. Available at https://goo.gl/oe2iVt [Accessed 25 November 2022]

(7) Haase J, Lavanderos S, Riquelme C, et al. Association of socio-demographic factors with the age at death due to cardiovascular diseases. Rev Med Chil. 2016;144(11):1464–1472. doi: 10.4067/S0034-98872016001100013

(8) Leslie KH, McCowan C, Pell JP. Adherence to cardiovascular medication: a review of systematic reviews. J Public Health (Oxf). 2019;41(1):84–94. doi: 10.1093/pubmed/fdy088

(9) Liu M, Zheng G, Cao X, et al. Better Medications Adherence Lowers Cardiovascular Events, Stroke, and All-Cause Mortality Risk: A Dose-Response Meta-Analysis. J Cardiovasc Dev Dis. 2021;8(11):146. doi: 10.3390/jcdd8110146.

(10) Bosomworth NJ. Practical use of the Framingham risk score in primary prevention: Canadian perspective. Can Fam Physician. 2011;57(4):417–23. PMCID: PMC3076470

(11) Chowdhury R, Khan H, Heydon E, et al. Adherence to cardiovascular therapy: a metaanalysis of prevalence and clinical consequences. Eur Heart J. 2013;34(38):2940–8. doi: 10.1093/eurheartj/eht295

(12) Ordunez P, Campbell NRC, Giraldo Arcila GP, et al. HEARTS in the Americas: innovations for improving hypertension and cardiovascular disease risk management in primary care. Rev Panam Salud Publica. 2022; 16;46: 96. doi: 10.26633/RPSP.2022.96

(13) Macinko J, Andrade FCD, Nunes BP, et al. Primary care and multimorbidity in six Latin American and Caribbean countries. Rev Panam Salud Publica. 2019;43: e8. doi: 10.26633/RPSP.2019.8. PMID: 31093232.

(14) World Health Organization. Sabaté E. Adherence to long-term therapies: evidence for action. Geneva: World Health Organization. 2003. Available at https://www.who.int/chp/knowledge/publications/adherence_full_report.pdf. [Accessed 7 December 2022]

(15) Kunstmann S, Lira MT, Molina JC, et al. Riesgo de Presentar un Evento Cardiovascular a 10 años en Personas Sanas: Proyecto RICAR. Rev. Chil Cardiol. 2004; 23:13–20.

(16) )Ritchie J, Lewis J. Qualitative Research Practice—A Guide for Social Science Students and Researchers. (2003). Available at https://www.scirp.org/(S(351jmbntvnsjt1aadkposzje))/reference/ReferencesPapers.aspx?ReferenceID=1401383 [Accessed 25 November 2022]

(17) Pope C, Ziebland S, Mays N. Qualitative research in health care. Analysing qualitative data. BMJ. 2000;320(7227):114–6. doi: 10.1136/bmj.320.7227.114

(18) Gale NK, Heath G, Cameron E, et al. Using the framework method for the analysis of qualitative data in multi-disciplinary health research. BMC Med Res Methodol. 2013; 13 :117. doi: 10.1186/1471-2288-13-117

(19) Kripalani S, Goggins K, Nwosu S, et al. Vanderbilt Inpatient Cohort Study. Medication Nonadherence Before Hospitalization for Acute Cardiac Events. J Health Commun. 2015;20 Suppl 2(0):34–42. doi: 10.1080/10810730.2015.1080331

(20) González-Bueno J, Calvo-Cidoncha E, Sevilla-Sánchez D, et al. Spanish translation and cross-cultural adaptation of the ARMS-scale for measuring medication adherence in polypathological patients. Aten Primaria. 2017;49(8):459–464. doi: 10.1016/j.aprim.2016.11.008

(21) Kripalani S, Risser J, Gatti ME, et al. Development and evaluation of the Adherence to Refills and Medications Scale (ARMS) among low-literacy patients with chronic disease. Value Health. 2009;12(1):118–23. doi: 10.1111/j.1524-4733.2008.00400.x

(22) Lomper K, Chabowski M, Chudiak A, et al. Psychometric evaluation of the Polish version of the Adherence to Refills and Medications Scale (ARMS) in adults with hypertension. Patient Prefer Adherence. 2018; 12 :2661–2670. doi: 10.2147/PPA.S185305

(23) Bosanquet K, Bailey D, Gilbody S, et al. Diagnostic accuracy of the Whooley questions for the identification of depression: a diagnostic meta-analysis. BMJ Open. 2015;5(12): e008913xs. doi: 10.1136/bmjopen-2015-008913

(24) Bradley KA, DeBenedetti AF, Volk RJ, et al. AUDIT-C as a brief screen for alcohol misuse in primary care. Alcohol Clin Exp Res. 2007;31(7):1208–17. doi: 10.1111/j.1530-0277.2007.00403.x

(25) Navickas R, Petric VK, Feigl AB, et al. Multimorbidity: What do we know? What should we do? J Comorb. 2016;6(1):4–11. doi: 10.15256/joc.2016.6.72

(26) Masnoon N, Shakib S, Kalisch-Ellett L, et al. What is polypharmacy? A systematic review of definitions. BMC Geriatr. 2017;17(1):230. doi: 10.1186/s12877-017-0621-2

(27) Gast, A., Mathes, T. Medication adherence influencing factors—an (updated) overview of systematic reviews. Syst Rev. 2019; 8(1):112. doi: 10.1186/s13643-019-1014-8

(28) Lee H, Park JH, Floyd JS, et al. Combined Effect of Income and Medication Adherence on Mortality in Newly Treated Hypertension: Nationwide Study of 16 Million Person-Years. J Am Heart Assoc. 2019;8(16): e013148. doi: 10.1161/JAHA.119.013148

(29) Crawshaw J, Auyeung V, Norton S, et al. Identifying psychosocial predictors of medication non-adherence following acute coronary syndrome: A systematic review and meta-analysis. J Psychosom Res. 2016; 90 :10–32. doi: 10.1016/j.jpsychores.2016.09.003

(30) Alsabbagh MH, Lemstra M, Eurich D, et al. Socioeconomic status and nonadherence to antihypertensive drugs: a systematic review and meta-analysis. Value Health. 2014;17(2):288–96. doi: 10.1016/j.jval.2013.11.011

(31) Lemstra M, Alsabbagh MW. Proportion and risk indicators of nonadherence to antihypertensive therapy: a meta-analysis. Patient Prefer Adherence. 2014; 8 :211–8. doi: 10.2147/PPA.S55382

(32) Rasmussen JN, Chong A, Alter DA. Relationship between adherence to evidence-based pharmacotherapy and long-term mortality after acute myocardial infarction. JAMA. 2007;297(2):177–86. doi: 10.1001/jama.297.2.177

(33) Khatib R, Schwalm JD, Yusuf S, et al. Patient and healthcare provider barriers to hypertension awareness, treatment and follow up: a systematic review and metaanalysis of qualitative and quantitative studies. PLoS One. 2014;9(1): e84238. doi: 10.1371/journal.pone.0084238

(34) Wei J, Hou R, Zhang X, et al. The association of late-life depression with all-cause and cardiovascular mortality among community-dwelling older adults: systematic review and meta-analysis. Br J Psychiatry. 2019;215(2):449–455. doi: 10.1192/bjp.2019.74

(35) Puschel K., Furlan E., Dekkers W. Social Health Disparities in Clinical Care: A New Approach to Medical Fairness. Public Health Ethics. 2015; 10, 78–85. doi: 10.1093/phe/phv034

(36) Engberg I, Segerstedt J, Waller G, et al. Fatigue in the general population-associations to age, sex, socioeconomic status, physical activity, sitting time and self-rated health: the northern Sweden MONICA study 2014. BMC Public Health. 2017;17(1):654. doi: 10.1186/s12889-017-4623-y

(37) Vaes AW, Goërtz YMJ, van Herck M, et al. Physical and mental fatigue in people with non-communicable chronic diseases. Ann Med. 2022;54(1):2522–2534. doi: 10.1080/07853890.2022.2122553

(38) García-Huidobro D, Bittner M, Brahm P, et al. Family intervention to control type 2 diabetes: a controlled clinical trial. Fam Pract. 2011; 28(1):4–11. doi: 10.1093/fampra/cmq069

(39) Jeemon P, Harikrishnan S, Ganapathi S, et al. Efficacy of a family-based cardiovascular risk reduction intervention in individuals with a family history of premature coronary heart disease in India (PROLIFIC): an open-label, single-centre, cluster randomised controlled trial. Lancet Glob Health. 2021;9(10): 1442–1450. doi: 10.1016/S2214-109X(21)00319-3

